# Predictive Modelling of Brain Disorders with Magnetic Resonance Imaging: A Systematic Review of Modelling Practices, Transparency, and Interpretability in the use of Convolutional Neural Networks

**DOI:** 10.1101/2021.11.20.21266620

**Authors:** Shane O’Connell, Dara M Cannon, Pilib Ó Broin

**Author notes:** Provided equal supervision (Dara M Cannon, Pilib Ó Broin).

## Abstract

Brain disorders comprise several psychiatric and neurological disorders which can be characterised by impaired cognition, mood alteration, psychosis, depressive episodes, and neurodegeneration. Clinical diagnoses primarily rely on a combination of life history information and questionnaires, with a distinct lack of discriminative biomarkers in use for psychiatric disorders. Given that symptoms across brain conditions are associated with functional alterations of cognitive and emotional processes, which can correlate with anatomical variation, structural magnetic resonance imaging (MRI) data of the brain are an important focus of research studies, particularly for predictive modelling. With the advent of large MRI data consortia (such as the Alzheimer’s Disease Neuroimaging Initiative) facilitating a greater number of MRI-based classification studies, convolutional neural networks (CNNs) – deep learning models suited to image processing – have become increasingly popular for research into brain conditions. This has resulted in a myriad of studies reporting impressive predictive performances, demonstrating the potential clinical value of deep learning systems. However, modelling practices, transparency, and interpretability vary widely across studies, making them difficult to compare and/or reproduce, thus potentially limiting clinical applications. Here, we conduct a qualitative systematic literature review of 60 studies carrying out CNN-based predictive modelling of brain disorders using MRI data and evaluate them based on three principles – modelling practices, transparency, and interpretability. We furthermore propose several recommendations aimed at maximising the potential for the integration of CNNs into clinical frameworks.

## 1 Introduction

Brain disorders, which include bipolar disorder, alzheimer’s disease, and schizophrenia, are a collection of debilitating neurological and psychiatric conditions characterised by a variety of features including impaired cognition, altered mood states, psychosis, neurodegeneration, and memory loss [1]. These phenotypes, each with varied clinical presentations, are all associated with neuroanatomical changes, incurring public and personal health burdens through reduced quality of life, social stigma, and increased mortality [1, 2]. As such, these conditions are the focus of intense research across multiple disciplines. There is significant interest in building predictive models designed to differentiate conditions and their subtypes, which could incorporate biological information into current clinical frameworks and yield mechanistic insights via the biomarkers used [3, 4]. Additionally, biomarker-informed diagnoses could offer the potential of early intervention and management, a concept well understood in general medicine [5]. Magnetic resonance imaging (MRI) provides non-invasive measures of brain structure and the increasing availability of large-scale collections of MRI data has enabled a wealth of predictive modelling studies [6, 7].

Previously, machine learning and classical statistical approaches have been used to highlight differential neuroanatomical patterns across several conditions, including subcortical structure volume reduction in bipolar disorder and alzheimer’s disease [8, 9]. However, incorporating such information into clinical systems is non-trivial, as the dynamics and limitations of a particular biomarker must be addressed prior to use [10, 11]. Additionally, the methods used to identify discriminative features have their own considerations, such as requiring preprocessing tools to derive tabular brain summary information [12, 13]. These tools can produce variable results depending on the parameters chosen, even when applied to the same dataset, highlighting the importance of domain expertise to justify decisions [14]. Additionally, statistical modelling often requires formal specification of expected variable relationships, and generally are unsuited to high-dimensional imaging data structures. Traditional machine learning approaches are also limited by their inability to consider spatial dependencies between groups of pixels, making it necessary to use tabular summary data. With these factors in mind, deep learning algorithms – and particularly those well-suited to imaging – have become a popular methodology. This is because of their ability to consider arbitrarily complex relationships, meaning greater model flexibility without specification of expected variable relationships. Convolutional neural networks (CNNs) are deep learning models designed to detect spatial patterns in imageing data and have shown impressive predictive performances in various classification tasks. They have also been widely applied in the field of medical imaging for segmentation and prediction, particularly in the context of aging and psychiatric/neurological disorder diagnosis [15, 16, 17, 18, 19, 20, 21, 22].

These recent developments have been enabled by access to large standardised neuroimaging data collections, such as the alzheimer’s Disease Neuroimaging Initiative [23] and the UK Biobank [24]. The predictive capabilities of these approaches is promising in the context of potential clinical applications; brain disorder classifications are usually based on life history information and questionnaires, and thus leveraging models making use of neuroanatomical measures could supplement existing diagnostic frameworks. However, there are a few caveats that bear consideration; firstly, deep learning models have a number of limitations, such as high parameter dimensionality, lack of interpretability, weight stochasticity, lack of uncertainty, and difficulty to train [25, 26, 22, 27]. Secondly, clinical decision systems require rigorous validation and reporting frameworks for more interpretable models; the use of opaque deep learning algorithms make validation and transparency more difficult to achieve [28, 29]. Clinical decision systems that offer no explanation of a classification are less likely to be incorporated into patient care frameworks. These factors combine to make the application of deep learning to clinical settings challenging, particularly where medical imaging is concerned.

As the number of studies applying deep learning to brain disorder prediction using neuroimaging data increases, the opportunity arises to examine factors that may limit their potential for clinical application. In this work, we systematically review 60 papers which report on such approaches. While many of the studies examined have been designed to demonstrate the predictive capabilities, we sought to assess the existing literature with the aim of identifying key principles that can maximise the potential clinical value of future work; these principles are: 1) modelling practices, 2) transparency, and 3) interpretability. Below, we first provide a brief overview of CNNs and their workflow in the context of brain disorder imaging-based models, and subsequently detail our motivation behind these three principles; we then analyse the selected papers in the context of these principles and suggest several recommendations for future studies based on our results.

### 1.1 Convolutional Neural Networks

CNNs are a popular deep learning algorithm for many areas of research, particularly those utilising MRI data [15, 16, 17, 18, 19, 20, 21]. Their structure is designed to account for spatial data patterns; this is accomplished through the use of filters and feature maps. A feature map is derived via *convolutional operations*, which are a matrix multiplication between a weights vector of an arbitrary size (the filter, for example, may be 2 *×* 2 pixels large in a 2D example) and an input image patch of the same size. The result, which is every number in the input multiplied by every number in the filter and summed together, is then the pixel of a new feature map. The convolution of the same filter over every patch of the input image generates the entire output feature map, which is usually the same size as the input image. Multiple feature maps are used in CNN architectures, each with their own filters, which, throughout model training, can detect distinct data patterns such as shapes and/or edges. The goal of CNNs is to build increasingly abstract representations of data through iterative downsampling and transformations until such a time as linear seperation of classes is possible in predictive tasks. Weight initilisation is often random and training is carried out via backpropagation. More in-depth considerations of neural networks and their training can be found in LeCun et al., 1995 and 2012 [22, 27].

### 1.2 CNN Implementations

MRI-based predictive modelling of brain conditions with deep learning models generally follows the pipeline presented in Figure 1, or a variant thereof. Preprocessing is usually applied to skull strip, register raw input images, crop, resize, and/or contrast normalise. The preprocessed inputs are then used as training data for a CNN (or an ensemble of CNNs). Owing to the fact that many existing CNN models have been applied to 2D data domains, studies in the medical imaging field can adapt their data to fit existing architectures via transfer learning or train new models in the 3D space, as structural MRI scans are usually 3D [30, 31]. Some studies also train custom architectures on 2D data [32, 33, 34]. The output is usually presented as a probability, which is then used to calculate performance metrics such as the area under the receiver operating characteristic curve (AUC) and accuracy.

**Figure 1:**
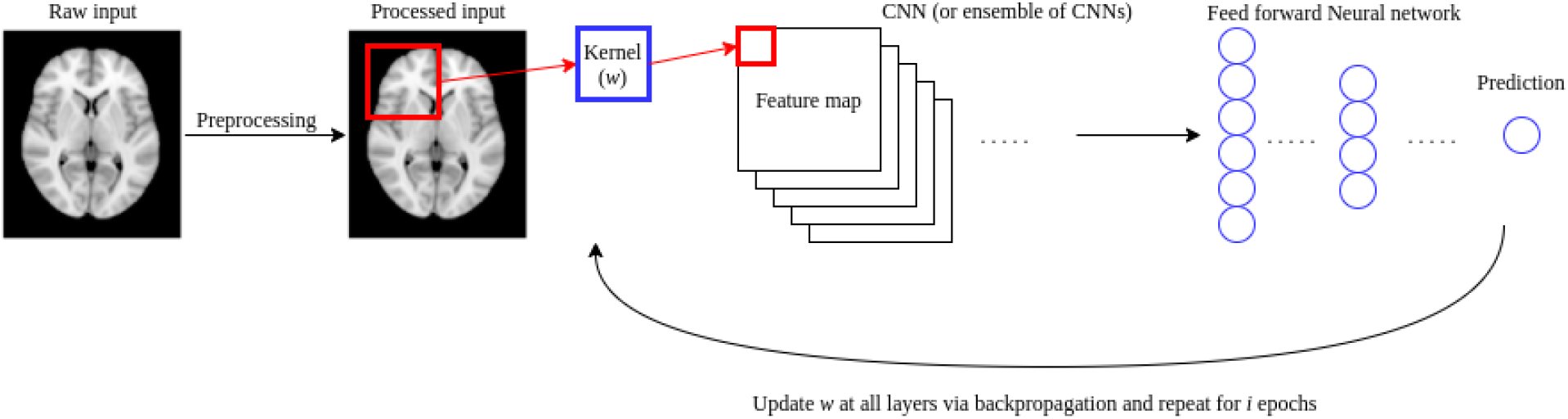
General experimental workflow. The preprocessed input image, either in 2 or 3 dimensional format is passed to a CNN model (or ensemble of CNN models) for training and prediction, The weights vector w is updated via backpropagation at each epoch, minimising the error of the loss function.

In the following sections, we define and justify our emphasis on modelling practices, transparency, and interpretability in the context of brain disorder classification using neuroimaging data for potential clinical benefit. We note that these principles bear domain-agnostic importance for predictive studies and overlap with recent recommendations for improving the clinical and biological translational potential of machine learning and deep-learning models [35].

### 1.3 Modelling Practices

Modelling practices here refers to the reliability of the methodology used; methodologies optimising reproducibility efforts via approaches demonstrating the reliability of experimental results are increasing the potential clinical value of a study. Generally, studies that can be reproduced and that have attempted to mitigate factors that can influence the reliability of results are more likely to witness clinical integration. As previously mentioned, deep learning models have a number of unique features that make this task difficult, but several procedures can be observed. We examine the presence of repeat experiments, the data splitting procedure, potential information leakage, and the data representation strategy to evaluate this principle. Repeat experiments ensure that the reported performance metrics are trustworthy across multiple random weight initialisations and that the system as a whole can be expected to perform well if retrained. This is pertinent given that CNNs are paramater-dense, making them more prone to overfitting. A useful type of repeat experiment includes *k*-fold cross validation, whereby data is split into *k* folds, and *k* − 1 folds are used to train the model and the *k −*1 fold serves as the testing set. This procedure is repeated *k* times, until every fold has served as the testing set, providing an estimate of model performance on multiple data splits.

The data splitting procedure is important as training/testing separation must be ensured for robust performance estimation. Information leakage describes situations whereby model testing and training sets are not kept entirely independent during model tuning, which may lead to inflated performance metrics; this can occur where model performance, usually estimated from entirely independent data, is partly estimated on examples already used to optimise weights. This can limit reproducibility and ultimately the potential clinical value of an approach.

The data representation strategy is of specific importance for CNN models in this domain, as structural MRI data is 3D, whereby each number is represented by a pixel. Thus, modelling entire volumes can be computationally expensive, and some studies may opt to split data into individual 2D slices. This comes with a set of caveats: firstly, each 2D slice is treated as an individual instance during conventional training procedures, meaning that performance metrics can either be reported per slice or combined to derive patient-level quantities, prompting consideration of voting strategies; secondly, 2D data are more prone to information leakage if train-test splitting is carried out after 2D slice derivation. Together, these issues can contribute to inflated estimates of performance.

### 1.4 Transparency

Transparency refers to how clearly the study’s methods are reported, including code and model sharing. This principle bears general importance, particularly for models with clinical potential [36]. Several important advantages to code sharing having been described previously, including facilitating greater understanding of experiments and facilitating reproducibility [37, 38]. There are many hyperparameters associated with deep learning models which can effect performance, making transparent reportage necessary. Descriptions of model architectural choices and training schedules can help to increase potential for clinical translation through increased reproducibility and understanding of studies. Furthermore, model weight sharing can mitigate the computational overhead of model training.

### 1.5 Interpretability

Interpretability refers to the efforts made to explain features driving model predictions. Deep learning systems are not well suited to interpretation, but efforts can be made to examine image regions that are used during prediction to determine whether that information is relevant. This is particularly important as CNNs are prone to overfitting and can make use of any image feature, in turn making algorithmic biases more likely if not examined [39, 40]. Ensuring CNNs are using relevant information can increase clinical potential and confidence in the system. Models can be interpreted by saliency methods such as gradient-based class activation mapping [41, 17], which rely on deriving the gradient of model output with respect to input and weighting that quantity by the input – the final metric is then overlaid on the input for visualisation. This can indicate what regions are most ‘important‘, but they do not offer the same explanatory power as coefficients from regression models. Another approach to understanding model behaviour is counterfactuals, which involve measuring the changes in predictive performance of models when they are exposed to inputs with known qualities; for example, noting the change in model output when a patient image with a thicker amygdala is used as the input [42].

## 2 Methods

We conducted a systematic literature review according to PRISMA guidelines [43], the details of which are provided below.

### 2.1 Inclusion/exclusion criteria

We limited our search to consider studies making use of CNNs exclusively (end-to-end), and convolutional layer outputs, or other model outputs, are not used to train separate machine learning models. This is because they are the most common architecture. We also focused our attention on studies that use structural MRI data, as functional MRI data structures can often have different modelling requirements, including the use of time series methodologies that make them more difficult to compare.

### 2.2 Search details

We performed a Web of Science (all databases) and Pubmed search with the following keywords:

((((structural) or (T1-weighted)) AND (imaging)) AND ((MRI) OR (T1 MRI)) AND ((CNN) OR (convolutional neural network) OR (3D-CNN))) AND (psychiatric OR depression OR autism OR bipolar OR Alzheimer’s OR neurological) NOT (segmentation)

For Web of Science, 77 results were returned, and 114 results were returned from Pubmed. Titles and abstracts were screened for relevance to the research question, and duplicates across both databases were removed, leaving a total of 76 papers. 16 studies were excluded for using functional MRI data and using hybrid models where CNNs were not the primary modelling method; this resulted in a total of 60 papers remaining for review. The flowchart of this process is presented in Figure 2.

**Figure 2:**
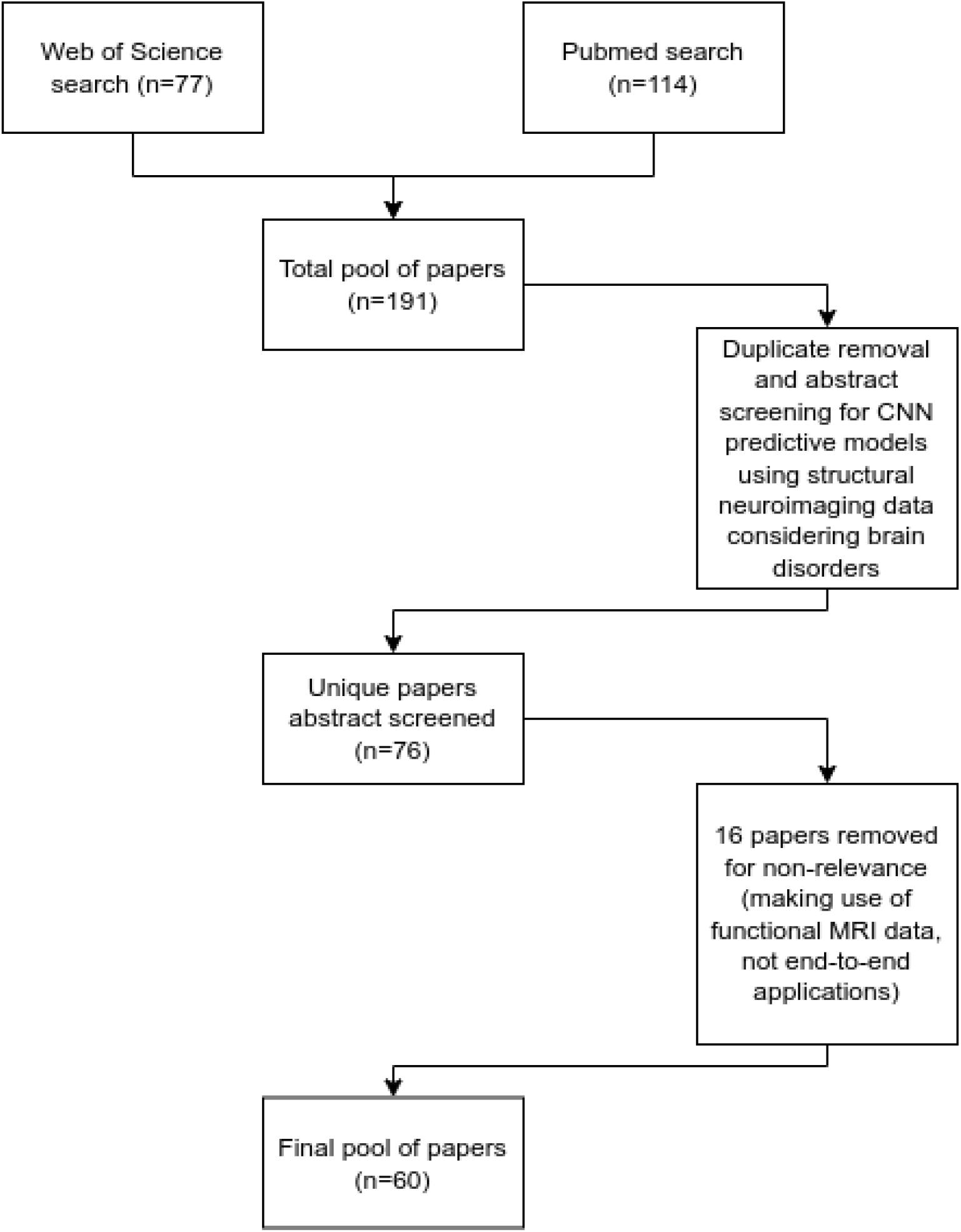
Flowchart detailing the paper selection process.

### 2.3 Desired variables

A standardised questionnaire was designed to evaluate the methodological details of the studies considered. No numerical variables were sought as this work aimed to qualitatively examine the outlined principles.

## 3 Results

We organise our findings according to our three principles: modelling practices, transparency, and interpretability. The selected papers and their attributes can be found in Table 1, and a numerical summary of the results can be found in Table 2.

**Table 1:**
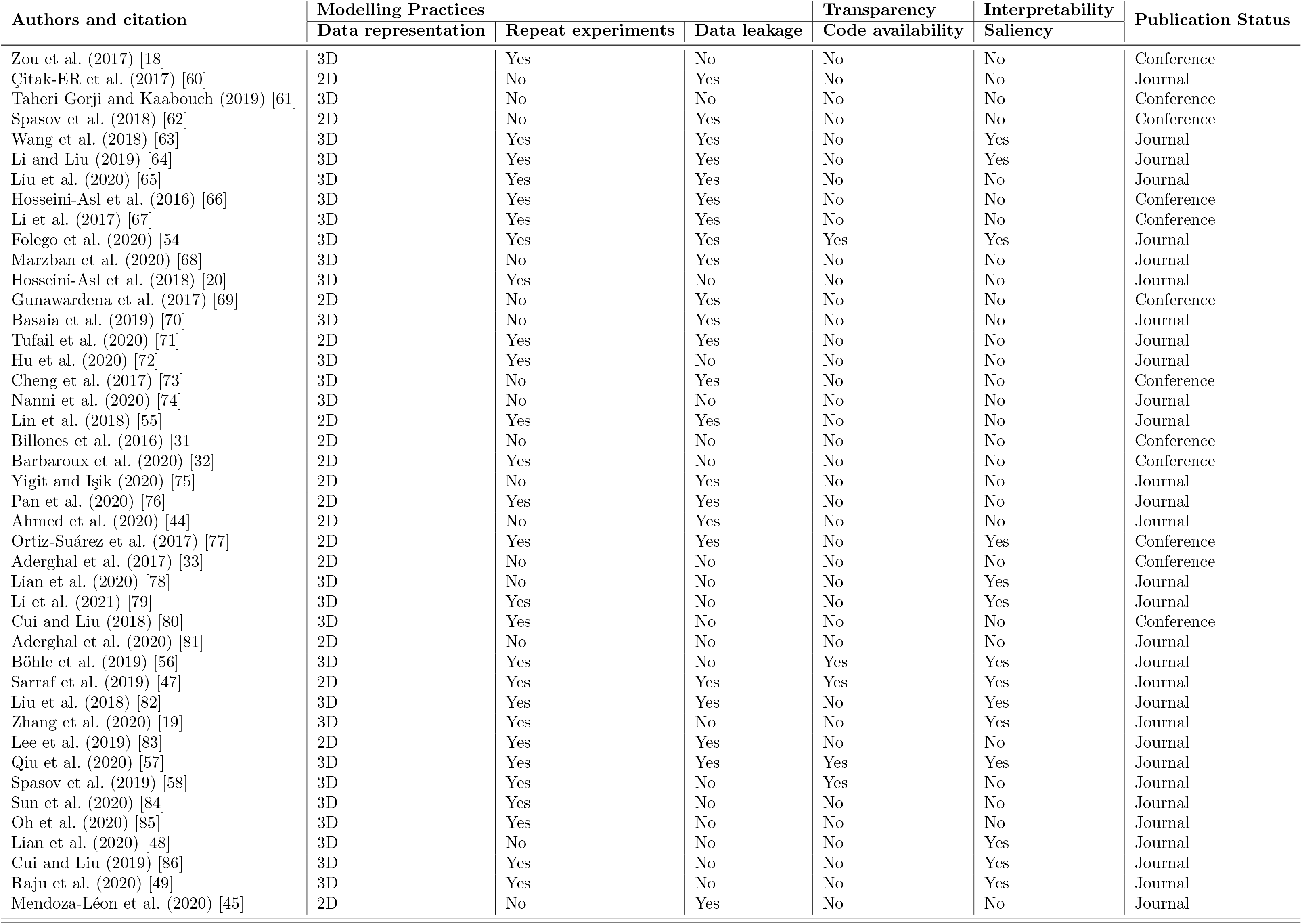
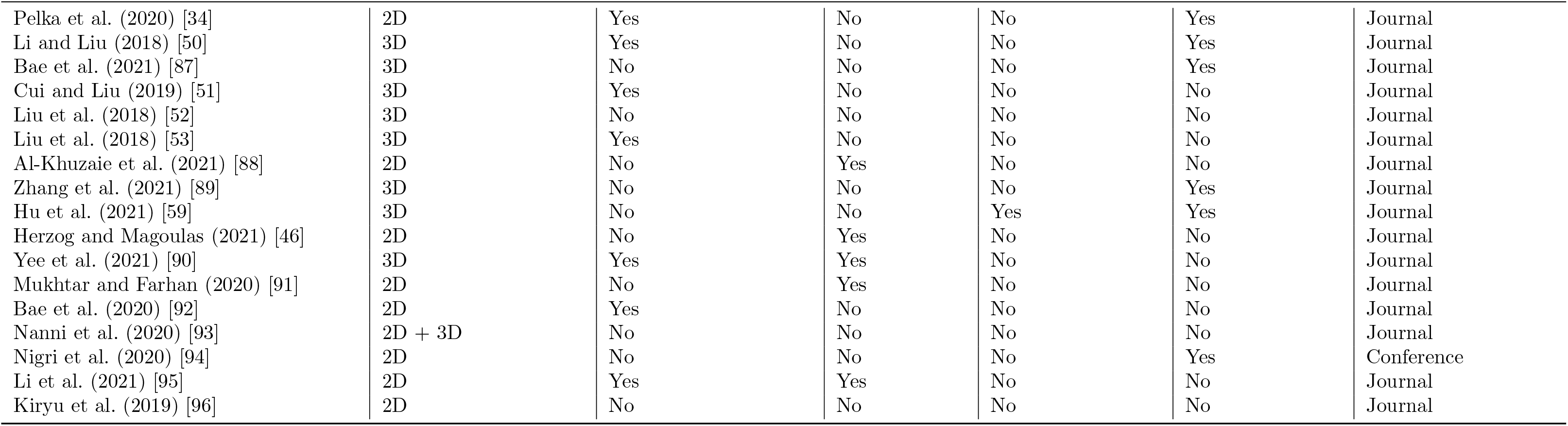
Tabular presentation of the studies considered for this systematic literature review.

**Table 2:** Numeric summary of study attributes from the 60 papers satisfying selection criteria.

| Question | Answer |
| --- | --- |
| How are data represented? | 2D(n=24),<br>3D(n=35),<br>Both(n=1) |
| Is code available? | No(n=54),<br>Yes(n=6) |
| Conference or Journal? | Conference(n=13),<br>Journal(n=47) |
| Is saliency considered? | No(n=40),<br>Yes(n=20) |
| Are there repeat experiments? | No(n=26),<br>Yes(n=34) |
| Is there potential information leakage? | No(n=32),<br>Yes(n=28) |

### 3.1 Modelling practices

We found that 24 out of 60 papers represented data in 2D (Table 2). While this is more computationally efficient than 3D, it can make information leakage more likely. Accuracy calculation can be carried out per slice or per patient, introducing issues surrounding optimal voting strategies. Of the 24 studies making use of 2D slices, only one referred to voting methods, and 15 studies suffered from potential leakage [44]. Leakage was flagged where it was clear that training/test splitting occurred per slice and not per patient or where it could not be ruled out based on methodological descriptions. Several studies made use of single slices per patients [45, 46, 33]. Even with registration, there is no guarantee that the same biological information is considered per patient with this approach. Spatial dependencies between regions are ignored when using 2D data, which may have implications for mechanistic understanding and predictive capabilities. One paper making use of 2D slices provided code [47]. We noted that 25 out of 60 studies made use of multiple models for training and prediction, with some papers using the output of one trained CNN as the input to another [48, 49, 50, 51, 52, 53]. This may facilitate overfitting by impacting generalisation. A number of studies used statistical tests to pre-select informative image patches which can introduce bias by focusing the model on regions which may not be informative in full models. [53, 52, 45]. Furthermore, pre-selecting regions based on accuracy metrics in one population may influence generalisability in another, which is pertinent for clinical potential. In several studies, one model was trained on the whole dataset the weights from that model were used for transfer learning of another model, leading to potential leakage or overfitting [54, 55, 44, 45, 34].

Thirty four out of 60 studies employed repeat experiments. Ten such papers reported point estimates, and 5 provided code [56, 47, 57, 54, 58]. Of the 15 studies with repeat experiments and interpretability efforts, none detailed whether saliency methods were applied per fold or on test sets.

### 3.2 Transparency

We found that 54 out of 60 papers did not provide code or model weights, meaning that the majority of studies relied on textual methods summaries. This implies limited methodological transparency; this is an issue when considering how modelling choices can impact system performance. Studies reporting code facilitate clear, reproducible experimental practices [54, 56, 57, 58, 59, 47].

### 3.3 Interpretability

Twenty of 60 studies considered interpretability by applying a saliency method [41] or visualising feature maps [17]. Of these 20, 5 papers discussed their interpretation of saliency outputs in their findings, with the remainder providing little to no commentary [56, 82, 57, 49, 94]. Four such papers with discussions made use of single saliency methods, with two out of five providing code [56, 57]. Nine studies presented interpretability results with little to no commentary and no code [64, 65, 78, 79, 47, 82, 86, 50, 87]. Of the remaining 11 studies, 5 provided code, with a combination of saliency and transparency [56, 47, 57, 54, 59]. A subset of studies underscored that expert-driven preprocessing is not required with deep learning studies, and almost all studies alluded to this fact in their introductions [59, 68, 78, 50, 51].

## 4 Discussion

We conducted a systematic literature review of 60 studies carrying out CNN-based predictive modelling of brain disorders using MRI data and evaluated their modelling practices, transparency, and considerations of interpretability in the context of their potential clinical value. Our results identified several areas for potential improvement across three principles that we believe will maximise the potential for clinical integration. Below, we discuss the findings summarised in Table 2 in and propose several recommendations to maximise the potential clinical value of future studies.

### 4.1 Data representation

A majority of papers made use of 3D data representations, which is sensitive to aforementioned CNN-specific limitations of 2D data. This ensures that all biological information is used during training, as opposed to individual slices where spatial inter-dependencies are ignored. There was a significant minority of papers making use of 2D data structures (24/60), which may pose issues for downstream clinical applications. Two-dimensional models have multiple caveats discussed previously, including multiple majority voting strategies and potential information leakage. The high computational cost of modelling on 3D data structures impedes their implementation. Thus, leveraging 2D model weights for transfer learning is attractive, and we recommend cognizance of the limitations associated with 2D modelling and attempt to mitigate these issues. For example, researchers can examine how performance metrics change relative to different voting strategies. Regarding information leakage, researchers can ensure slice conversion post-data splitting at the patient level; providing code can ensure supplement this via transparency. Utilising single slices may lead to performance estimation inflation owing to the fact that there is no guarantee the same biological information is being considered per patient. Several studies also made use of model stacking, whereby the input of a model is the output of another trained model. This may impact the model’s ability to generalise to different data by increasing the chances of overfitting. This is because the first model in stacking situations has already derived a representation of the data, meaning the next model’s internal representation of the problem domain is built upon a an initial abstraction of the input data. This is distinct from using traditional dimensionality reduction techniques before training a predictive model in two ways – firstly, model stacking in this domain is often not unsupervised step and hence the first model’s representation is based upon knowledge of test labels. Secondly, deep learning systems are opaque, making it difficult to understand both the predictive model’s data representation and the specifics of the reduced dimensionality space used as the input for the predictive model.

### 4.2 Repeat experiments

Most studies implemented repeat experiments, which demonstrate robust modelling practices. Such procedures account for weight initialisation stochasticity and potential performance inflation arising from fold splitting. Averaging over multiple random weight start points can return accurate performance estimates, with cross validation schemes being a reliable model diagnostic. Twenty-six of the 60 considered papers did not employ repeat experiments, which reduces confidence in reported results. A number of repeat experiment studies reported point estimates, which does not convey the spread of performance variability. Code inaccessibility exacerbates this issue, leaving the reader unclear as to the procedure followed. Reporting an average as a point estimate versus the range may cause the reader to assume the range was not large when performance may have been variable across repeats. We recommend that researchers employ repeat experiments and report their results with means and standard deviations.

### 4.3 Code availability

Most studies did not provide code. Wen et al. (2020) [97] underlined the importance of fairness, accountability, and transparency in deep learning modelling studies, and code inaccessibility runs contrary to these principles. The construction of deep learning systems requires many algorithmic decisions which can influence performance, introduce bias, and impact reproducibility. Deep learning models optimise an objective function over a set of arguments, meaning that any decisions taken in preprocessing and model construction can affect the capabilities of the system as a whole, and propagate subjective choices throughout ostensibly objective models [40]. For instance, several studies have examined algorithmic biases against underrepresented and/or marginalised groups [98, 99, 100]. Aside from domain-specific benefits to code sharing, the larger scientific community has recently shifted towards open science frameworks, with several high-profile journals requiring methodological transparency [101, 102, 103, 37]. Therefore, we believe that code availability and transparent methodological descriptions are an important aspect of deep learning experiments in this domain independent of potential clinical applications. Within a patient-care context, we underscore the importance of constructing reproducible systems to increase trust, both from a clinician and patient perspective. Studies making code available are proactively embracing these essential principles. We further encourage that minimal Jupyter/Google Colab notebooks, and other literate programming tools, be explored to enhance understanding and reproducibility [104, 105, 37]. This would also have the useful properties of allowing researchers to examine pipelines and identify potential ‘blind spots’ that the model authors may have overlooked in their modelling decisions, encouraging accountability [97]. Additionally, model training is incredibly computationally intensive; having access to models trained in similar domains could enable transfer learning approaches and mitigate data representation issues. Therefore, we recommend that authors share model weights and code to increase the potential of clinical translation.

### 4.4 Saliency and interpretability

We found many studies did not interrogate their presented models to ensure that relevant information is being used. Where irrelevant information is included, such as skull thickness when examining alzheimer’s disease neurodegeneration, without confirmation that the model is utilising brain information, attempts at patient care integration will have limited success. Even in cases where known irrelevant information can be removed by preprocessing, visual maps can draw attention to previously-unknown irrelevant information. Opaque black box models are less likely to be implemented in patient care settings, making the use of interpretability efforts of crucial importance. As previously stated, algorithmic biases in predictive settings is concerning, and saliency methods can help researchers to identify sources of bias where they occur. Twenty studies investigated neuroanatomical features driving model predictions via saliency methods, thus demonstrating relevant information is used during model training. While there exists variability in application, especially in terms of region understanding, we nonetheless recommend that future studies apply saliency methods to highlight brain regions being used. Saliency experiments leveraging multiple methodologies and those offering transparency through code availability can further increase potential clinical utility.

However, interpretability methods have a number of limitations that may hamper their application, prompting a discussion around how best to understand opaque models. Most existing methods return an ‘importance’ per pixel, which has no direct link to human-interpretable neuroanatomy. Usually, it represents the degree of change in the output relative to a small perturbation in the input pixel value, collapsing a potentially non-linear relationship to single values. While it provides an empirical assessment of captured patterns, it offers little interpretative value compared to coefficients returned by classical statistics. The deep learning field in general is focused on prediction as opposed to inference, meaning that the mechanistic understanding of relationship dynamics is often secondary to test accuracy. This is challenging in the context of discovery and clinical settings. Furthermore, saliency methods have their own limitations arising from their algorithmic derivation of ‘importance’, which can effect interpretation [106]. Similarly, counterfactuals, while promising, are difficult to empiricise and require significant computational overhead. Nonetheless, interpretability efforts allow researchers to visually evaluate model attention, which, for clinical translation, can serve to increase confidence and reduce bias, a topic of concern with respect to models applied to society at large [99, 40].

These methods may also be used to generate new hypotheses for downstream experiments. Highlighting neuroanatomical regions discriminative for particular conditions may suggest they have mechanistic relevance. Biomarker categorisation is an important step towards expanding current clinical care practices.

### 4.5 Future perspectives and commentary

This systematic literature review highlights areas of focus across modelling practices, transparency, and interpretability in the context of maximising the potential for clinical utility. These points underscore long-standing differences between deep learning and classical statistics, whereby the former is usually concerned with predictive performance and the latter with making inferential statements. The predictive imperative has led to numerous advances in image processing, with several state-of-the-art approaches developed to address tasks not suited to classical statistics [17, 22]. Neural networks have clear advantages where inferential dynamics are not a concern.

However, as deep learning becomes more readily applied to medical imaging domains, with potential consequences for patients, dichotomies of prediction versus inference should be retired. Researchers can maximise potential clinical benefit and potentially increase the quality of patient care by embracing the principles of reproducibility, transparency, and interpretability for predictive models. This can increase the confidence in such methods and consequently increase the potential for future clinical integration. We summarise our key recommendations in Table 3.

**Table 3:** Key recommendations arising from the results of this systematic literature review, their benefits, and the risks associated with non-adherence.

| Key Recommendations | Benefits | Risk(s) mitigated |
| --- | --- | --- |
| Make well-annotated code freely available | - Improve chances of reproducibility | - Limit reproducibility efforts<br>- Models remain opaque |
|  | - Readers can better understand workflow |  |
|  | - Encourage accountability and transparency |  |
| Employ repeat experiments | - Improve confidence in model estimation | - Risk reporting overfitted results |
|  | - Mitigate random weight initialisation | - Performance estimation inflation |
|  |  | - Diminished confidence in system overall |
| Use saliency metrics and counterfactuals | - Validate that model is using relevant information | - Models remain opaque |
|  | - Potential biomarker discovery | - Diminished confidence in system overall |
|  | - Improve confidence in system overall | - Unsure what information is being used by models |
| Avoid 2D data structures where possible | - Ensure spatial information between slices is considered | - Information leakage is more likely to occur |
|  | - Lessen chance of information leakage | - No guarantee spatial information between slices is considered |
|  | - No requirement of multiple voting strategies | - Must consider multiple voting strategies |

| Key Recommendations | Benefits | Risk(s) mitigated |
| --- | --- | --- |
| Make well-annotated code freely available | - Improve chances of reproducibility | - Limit reproducibility efforts<br>- Models remain opaque |
|  | - Readers can better understand workflow |  |
|  | - Encourage accountability and transparency |  |
| Employ repeat experiments | - Improve confidence in model estimation | - Risk reporting overfitted results |
|  | - Mitigate random weight initialisation | - Performance estimation inflation |
|  |  | - Diminished confidence in system overall |
| Use saliency metrics and counterfactuals | - Validate that model is using relevant information | - Models remain opaque |
|  | - Potential biomarker discovery | - Diminished confidence in system overall |
|  | - Improve confidence in system overall | - Unsure what information is being used by models |
| Avoid 2D data structures where possible | - Ensure spatial information between slices is considered | - Information leakage is more likely to occur |
|  | - Lessen chance of information leakage | - No guarantee spatial information between slices is considered |
|  | - No requirement of multiple voting strategies | - Must consider multiple voting strategies |

## 5 Limitations

This work reviewed studies from 2 database sources, but is not guaranteed to have evaluated all available relevant research. This study also did not undertake a quantitative review of reported accuracy metrics. This work also did not include considerations of studies making use of functional neuroimaging data, which contains a large corpus of research. We note that this review highlights issues with modelling 2D data structures; we acknowledge that this may not be feasible with respect to limited computational power. The extent of information leakage across these studies may be lower than reported; we classified a study as having potential information leakage where it was not possible to rule out its occurrence in studies that may have been prone (where data is not represented with one quantity per patient). Additionally, we acknowledge the drawbacks of saliency methods, whereby they primarily offer a visual check of model focus; nonetheless. Finally, we have endeavoured to ensure that our evaluation of studies is neither reflective of overall study quality nor reductive with respect to three nuanced principles; we acknowledge that the nature of the questionnaire results may present our findings as such – our binary descriptors are intended to serve as a vehicle to explore nuanced concepts.

## 6 Conclusion

In summation, we conducted a systematic literature review of 60 studies carrying out CNN-based predictive modelling of brain disorders using structural brain imaging data and evaluated them in the context of their modelling practices, transparency, and interpretability. We set forth recommendations that we believe will increase the future potential clinical value of deep learning systems in this domain. Careful consideration of these concepts can help to inform a clinical framework that can effectively incorporate deep learning into diagnostic and prognostic systems, enhancing our ability to improve patient care.

## Data Availability

All papers analysed in this review are available on either PubMed or Web of Science.

## 8 Acknowledgements

This work was conducted with the financial support of Science Foundation Ireland under Grant number 18/CRT/6214.

## 9 Data availability

All studies in this systematic literature review are accessible via PubMed and Web of Science.

